# An LLM enabled real-time estimation of seasonal influenza vaccine effectiveness from social media data

**DOI:** 10.64898/2026.08.28.26361670

**Authors:** Michael J. Pavia, Ivan Flores Amaro, Donfang Xu, Graciela Gonzalez-Hernandez, Matthew Scotch

**Affiliations:** Biodesign Center for Environmental Health Engineering, Arizona State University, Tempe AZ USA; Department of Computational Biomedicine, Cedars-Sinai Medical Center, Los Angeles, CA USA; College of Health Solutions, Arizona State University, Phoenix, AZ USA

## Abstract

Influenza vaccine effectiveness (VE) is estimated from a limited number of clinics using a test-negative design. These standard estimates face geographic, temporal, and operational constraints. Using Twitter/X data, we applied few-shot chain-of-thought prompting to identify self-reported vaccination status and influenza test results, then implemented a test-negative-like design to estimate VE. Our estimates fell within the range of interim reports and could complement current systems, improving feasibility, timeliness, and scalability.

## Main Text

Annual monitoring of influenza vaccine effectiveness (VE) is essential for guiding vaccination policy, informing clinical preparedness, and anticipating seasonal disease burden. Influenza causes millions of illnesses and billions of dollars in associated healthcare costs and productivity losses each year, making timely VE estimates a public health priority^1,2^. Influenza VE varies between seasons and populations because of both host and viral factors. Host-specific factors, including prior infection history, prior vaccination, and comorbidities, shape the immune response to vaccination^1^. In addition, viral antigenic drift and shift^3^ can reduce antigenic match between vaccine strains and circulating viruses.

In the United States, the Centers for Disease Control and Prevention (CDC) estimates influenza VE each season through the US Flu VE Network and related programs using a test-negative design (TND), which compares the odds of vaccination between laboratory-confirmed influenza cases and test-negative controls with acute respiratory illnesses^4^. TND is the standard method for VE estimation^4^, but it is sensitive to bias from data collection heterogeneity and inconsistent testing practices^5^. These networks enroll patients from a limited number of sites that do not capture the full geographic, demographic or clinical diversity of the broader population^6^. Inconsistent definitions of influenza “season”^7^ further reduce comparability across studies and, because vaccine uptake and influenza incidence^8^ vary over time, affect the estimates themselves. Interim influenza VE estimates are reported only later in the season^9^, limiting their use for real-time public health decisions. These limitations highlight the need for complementary data streams that improve both timeliness and population coverage^10,11^.

VE estimates from CDC surveillance studies, and other health agencies, serve as benchmarks for evaluating the plausibility and accuracy of alternative VE data streams. Social media data is available at a national scale and with near real time, offering the potential to extend geographic coverage and improve timeliness. Prior work has shown that large language models (LLM) and natural language processing methods can extract health related signals from social media and support epidemiologic analyses, including disease surveillance^11^ and the characterization of population level health patterns^10^. Building on the LLM pipeline developed by Xu et. al.,^12^, we evaluated whether a TND applied to self-reported posts on Twitter/X can recover VE estimates consistent with CDC and European clinical surveillance benchmarks^9,13–15^. The objective is not to replace established surveillance methods but to determine whether a pipeline based on social media data can produce plausible influenza VE estimates earlier and at a larger scale. Because the underlying posts are collected passively and at national scale, such a pipeline could update estimates as a season unfolds, ahead of the interim and end-of-season windows in which clinical estimates are released, and cover regions the sentinel sites do not.

To evaluate this approach, we applied a TND to self-reported posts on Twitter/X across two consecutive influenza seasons, using the LLM labeling pipeline described by Xu et. al.,^12^. For each season, we identified users who reported both an unambiguous influenza vaccination status and an unambiguous influenza test outcome, classifying those with positive test results as cases and negative test results as controls. The 2024-2025 influenza season (tweets posted 9 November 2024 – 19 April 2025) comprised 597 users (547 positive tests, 50 negative tests; 73 vaccinated). The 2025-2026 influenza season (tweets posted 1 October 2025 – 27 April 2026) comprised 1,911 users (1813 positive tests, 98 negative tests; 800 vaccinated). Details on data collection and labeling are described in the methods.

Seasonal influenza VE was 29% (95% CI: -69 to 67; p = 0.4) for 2024-2025 and 43% (95% CI: 14 to 62; p = 0.01) for 2025-2026. For 2024-2025, the estimate fell below the CDC outpatient point VE estimates reported for any influenza illness (33%; 95% CI: 24 to 41), A(H1N1)pdm09 (37%; 95% CI, 24%–48%), B/Victoria (40%; 95% CI, 12%–59%), but above for influenza A(H3N2) (27%; 95% CI, 14%–39%)^15^. The 2025–2026 estimate (43%) was slightly above the CDC interim outpatient VE estimates (22%–41%; US Flu VE: 22% in adults; VISION: 38% in children and 34% in adults; NVSN: 41% in children)^9^ and hospitalization VE estimates (30% in adults and 41% in children)^9^. These comparisons should be interpreted cautiously because the CDC values reflect interim seasonal data, whereas our estimate reflects the full season. When we restricted our analysis to the same surveillance period, VE was 41% (95% CI: 3 to 64; p = 0.03), which falls within the CDC reported estimates. Additionally, both the restricted and full season 2025-2026 estimates are consistent with early season estimates out of Europe^13,14^.

We recovered a statistically significant VE estimate that is within the expected clinical range from public social media text in the higher volume 2025-2026 season, but not in the smaller 2024-2025 season. The 2025-2026 result demonstrates feasibility of an LLM-enabled TND with sufficient tweet accrual; the 2024-2025 season did not replicate this performance and was limited by few self-reported negative tests. Cross-seasonal stability remains unestablished. Because posts accumulate in near real time, such a pipeline could also inform midseason public health messaging and vaccination campaigns when a usable signal is present, while laboratory confirmed clinical estimates continue to serve as the reference standard for policy.

To assess how much accumulated tweet volume was needed for cumulative estimates to enter the expected range, we calculated cumulative weekly VE estimates for each season (Figure 1). In the larger 2025-2026 tweet corpus, the cumulative VE estimate first entered and remained within the CDC interim benchmark range on 10 January 2026 (Week 1 of 2026; VE = 38%, 95% CI -8 to 65; p = 0.09), approximately nine weeks before the CDC interim report was published^9,13^, although it did not reach significance until five weeks later. This early in range agreement suggests that, given sufficient tweet volume, a social media TND can approximate clinical VE estimates ahead of formal interim releases. Additionally, in both seasons, weekly tweet volume tracked CDC clinical laboratory percent positivity (Figure 1). Positivity and tweet volume both peaked earlier in 2025-2026 (week ending 20 December 2025) than in 2024-2025(week ending 22 February 2025), and self-reported positive tests peaked in the week ending 10 January 2026, consistent with the earlier onset of the 2025-2026 season^9,13,14^.

**Figure 1.**
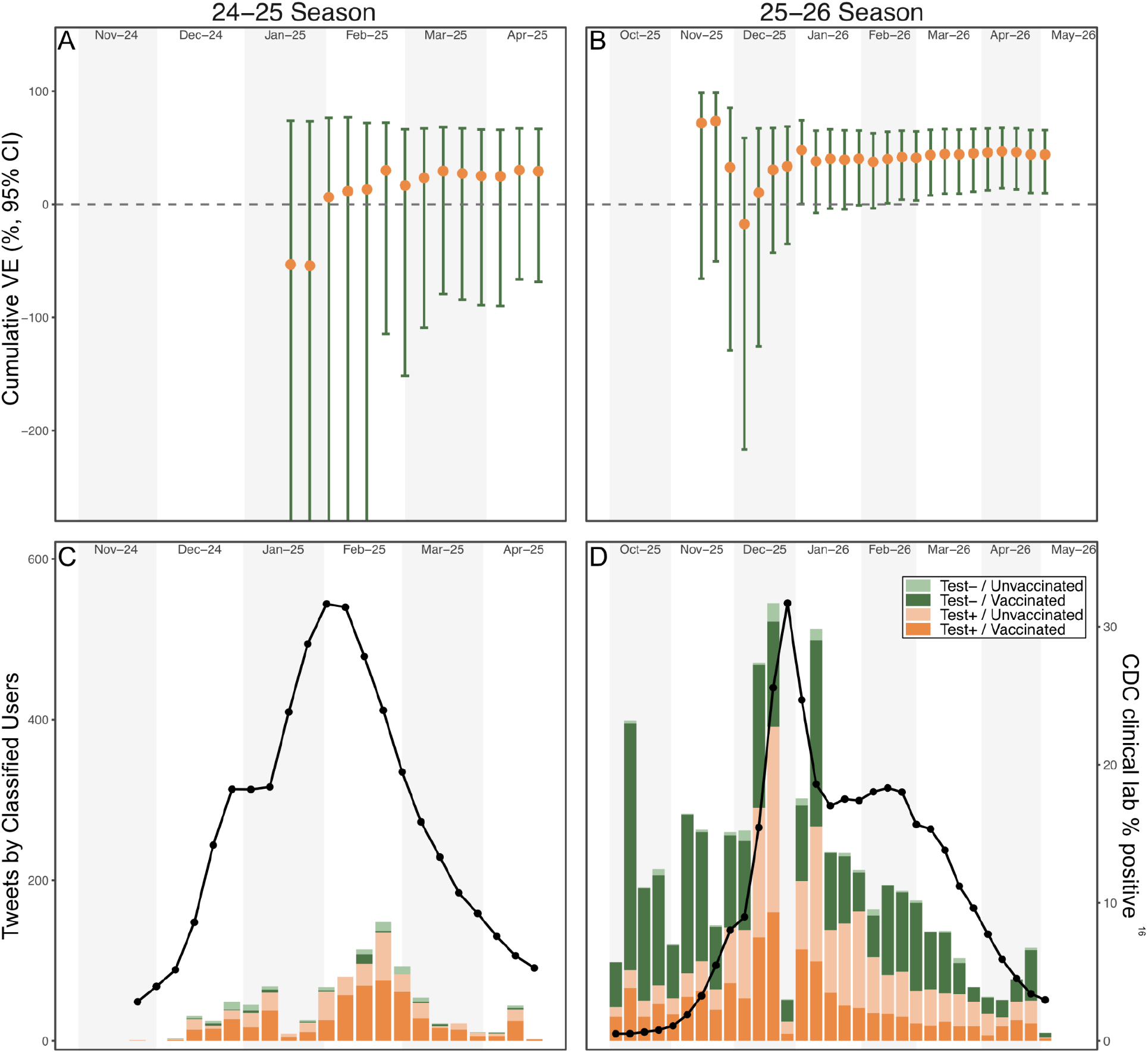
Influenza VE and Influenza-related Twitter/X activity during the 2024-2025 and 2025-2026 influenza seasons. (A, B) Weekly cumulative VE estimates for 2024-2025 (A) and 2025-2026 (B) influenza seasons, estimated using a TND from social media posts. Points indicate weekly VE estimates and error bars indicate 95% CI. The dashed horizontal line denotes VE = 0%. Estimates are shown only after at least 100 influenza positive cases were identified. (C, D) Weekly volume of user-linked tweets during 2024-2025 (C) and 2025-2026 (D) influenza seasons. Stacked bars indicated users by influenza test result and vaccination status. The black line shows the weekly percentage of positive influenza tests reported to CDC by clinical laboratories (right y-axis). CDC influenza surveillance data were obtained from FluView (https://www.cdc.gov/fluview/surveillance). In all panels the alternating shaded regions denote calendar months.

Our findings show that an LLM-enabled TND applied to public social media posts can recover a VE signal within the range of clinical TND, with potential to broaden participation and geographic reach. Several limitations qualify this result. The estimates are unadjusted as we did not collect patient age, comorbidities, or care-seeking behavior, which are routinely accounted for in clinical TND analyses^8^. In addition, inferred test and vaccination labels have some classification error (Xu et al.^12^ reported F1≥ 87% from the 2020-2021 season). However, this error is likely non-differential and would tend to underestimate the true VE^4^.

The 2024-2025 estimate was constrained by the small number of self-reported negative tests our sampling captured (50, against 547 positive tests), which produced a wide, non-significant confidence interval. The larger 2025-2026 cohort captured more negative tests (98) and yielded a precise, significant estimate despite a greater case-to-control imbalance (≈19:1 vs ≈11:1), indicating that the limiting factor was the number of controls rather than the ratio or a true scarcity of self-reported negative tests. Larger seasonal datasets or additional sampling would capture more controls and improve the precision of estimates. Finally, tweet timestamps are only approximate proxies for infection and vaccination dates and may affect within-season VE estimates.

Despite these limitations, an LLM pipeline applied to social media text produced statistically significant 2025-2026 VE estimates before the release of the interim CDC report. Our findings suggest that LLM-enabled TND analysis of social media may complement clinical VE surveillance. Used alongside established surveillance systems, this approach could extend geographic coverage, reduce reporting delay, and provide an early signal before clinical networks accrue sufficient enrollment for interim VE reporting.

## Methods

### Corpus collection and labeling

We retrieved tweets from the Twitter/X API using the keyword-plus-regular-expression collection pipeline outlined in Xu et al.,^12^. Broad keyword queries (e.g., “flu”, “influenza”) produced an initial corpus, which was refined with curated regular expressions capturing phrases indicative of flu testing (e.g., “had a positive flu test”) and flu vaccination (e.g. “got my flu shot”). Collection was done for both 2024-2025 and 2025-2026. For the 2024-2025 season we analyzed 42,994 tweets from 34,070 unique users posted between 9 November 2024 and 19 April 2025. For the 2025-2026 season we analyzed 199,054 flu test collection tweets from 136,058 unique users spanning CDC weeks ending 4 October 2025 to 2 May 2026. No identifiable personal information was collected and all downstream analyses used only the user identifier, tweet identifier, tweet text, and timestamp.

Each tweet was assigned a flu test outcome (positive, negative, other) or flu vaccination status (positive[current season], past, possible, negative, other) with the LLaMA-3-70B-Instruct using the few-shot chain-of-thought prompting framework of Xu et. al.,^12^ (inference via vLLM; temperature = 0.2, top-p = 0.2). The framework treats flu test outcomes and vaccination status as separate classification tasks defined by a fixed set of human annotation rules, with label definitions that separate a user’s current season experience from references to other people, prior seasons, stated intentions, or general flu discussion. Each prompt comprised task instruction, two randomly selected few-shot examples per label, and the target tweet with its posting date.

Vaccination status was assigned in a single classification step. Flu test outcome was assigned in two sequential steps: (1) first determine the test result (positive, negative, or other) without temporal information, and (2) determine whether the test occurred in the current season (yes, no, or other); the two outputs were integrated into the final flu test outcome label.

### Test-negative-like design cohort construction

For each season we constructed a user level TND cohort. Eligible users contributed at least one flu test labeled positive or negative and at least one flu vaccination labeled positive, past, or negative. When a user contributed multiple qualifying tweets, we retained the most recent flu test and the most recent flu vaccination status. A user was coded as vaccinated when the LLM labeled the vaccination tweet as “Positive” and that tweet was posted at least 14 days before the matched flu-test tweet; all other label combinations were coded as unvaccinated. Users with “Possible”, “Other” or missing labels were excluded.

### Vaccine effectiveness estimation

We estimated VE by logistic regression of case status on vaccination status. Exponentiated coefficients were taken as odds ratios, VE was defined as (1 − OR) × 100, and 95% confidence intervals were obtained by transforming profile-likelihood intervals on the OR^4,8^. To track VE as data accumulated over the season, we refit the model cumulatively by CDC week, including all users whose flu test tweet fell on or before each anchor week. Cumulative estimates were retained only when the window contained ≥100 test-positive users and both case and vaccination status had ≥2 levels; non-converging fits and those with non-finite confidence limits were discarded.

## Data Availability

All data produced in the present work are contained in the manuscript

## Data availability

Deidentified, user level summary datasets supporting the findings are available at https://github.com/pavia27/flu-ve-social-media-llm. Raw tweet text is not redistributed, consistent with the X Developer Agreement and Policy.

## Code availability

Analysis code reproducing all tables, VE estimates, sequential VE curves, and figures reported here is provided at https://github.com/pavia27/flu-ve-social-media-llm

## Acknowledgements

Research reported in this publication was supported by the National Library Of Medicine of the National Institutes of Health under Award Number R21LM01446 to MS. and GGH. The content is solely the responsibility of the authors and does not necessarily represent the official views of the National Institutes of Health.

## Author contributions

MJP, IFA, DX, GGH, and MS conceived and designed the study. IFA and DX collected and classified the tweet corpus. MJP analyzed the data. MJP wrote the original draft. MJP, IFA, DX, GGH, and MS reviewed and edited the paper. MS and GGH obtained funding. MS and GGH provided project administration.

